# Assessment of Awareness and Willingness to Use Pre-Exposure Prophylaxis for HIV Prevention among Female Sex Workers in Rwanda

**DOI:** 10.1101/2024.12.12.24318894

**Authors:** Albert Nzungize, Athanase Munyaneza

## Abstract

**Background:** Female sex workers (FSWs) in sub Saharan Africa bear a disproportionate burden of HIV infection. While pre exposure prophylaxis (PrEP) is increasingly accessible in the region, limited data exist on FSWs awareness of and willingness to use PrEP. This study aimed to assess PrEP awareness and willingness to use it, along with associated factors, among FSWs in Kigali, Rwanda.

**Methods:** This cross sectional study, conducted from April to September 2024, evaluated PrEP awareness and willingness to use it among 333 FSWs recruited through community based FSW associations. Data were analyzed using SPSS, with logistic regression employed to explore associations between participant characteristics and PrEP awareness and willingness. Ethical approval was obtained from the Rwanda National Ethics Committee.

**Results:** The mean age of participants was 30 years (SD: 7.26), with 47% aged between 25 and 34. Most were single (67%) and unemployed (66%), with nearly half (49.5%) reporting only primary education. A significant proportion (81%) had undergone HIV testing in the past six months, and 50% had been involved in sex work for 2 to 5 years. The median number of sexual partners in the past week was 7.

Awareness of PrEP was high (81%), however, consistent condom users were less likely to be aware of PrEP (adjusted odds ratio (aOR): 0.40, 95% confidence interval (CI): 0.19, 0.83), as were those not screened for sexually transmitted infections (STIs) compared to those who were screened (aOR: 0.43, 95% CI: 0.22, 0.85). Willingness to use PrEP was reported by 80% of participants. Those with a primary education were more willing to use PrEP compared to those with no formal education (aOR: 4.09, 95% CI: 1.62, 10.33). Conversely, participants screened for STIs were less likely to report willingness compared to those not screened (aOR: 0.28, 95% CI: 0.12, 0.62).

**Conclusion:** This study demonstrates high awareness and willingness to use PrEP among FSWs in Kigali. However, consistent condom users and those unscreened for STIs were less aware of PrEP, and willingness to use it varied based on education and STI screening status. These findings underscore the need for targeted health education and STI screening initiatives to enhance PrEP uptake and strengthen HIV prevention efforts in this vulnerable population.

## Background

HIV remains a critical public health issue, especially among key populations (KPs), such as female sex workers (FSWs). While the global median HIV prevalence among adults aged 15–49 years was 0.8%, it was significantly higher among FSWs, with a median prevalence of 3% in 2023(1). This highlights the elevated vulnerability of FSWs, with their HIV risk nearly four times that of the general adult population.

In sub-Saharan Africa (SSA), where the HIV prevalence is higher than that in any other global region, data from 2021 revealed that KPs and their sexual partners accounted for 51% of new HIV infections (2). The overall HIV prevalence among adults in Rwanda is 3%(3) and 4.3% in the city of Kigali(3) but is significantly higher among FSWs (51% to 57% in Kigali) (2). The annual HIV incidence is notably elevated for FSWs at 1.36%(4)(5)(6)

HIV pre-exposure prophylaxis (PrEP) can substantially decrease new HIV infections, with consistent adherence, reducing the risk of HIV acquisition by nearly 99% (7). Despite a willingness among FSWS in SSA to use PrEP(8) (9)(10)(11), multiple barriers impact access and adherence for these populations, including limited awareness of PrEP(8) (12), fear of side effects, social stigma from PrEP use, lack of permission or approval to use from client or partner, fear of perception regarding HIV-positive status (13), and difficulties with daily oral pill use (14) or sex work(15). Additionally, social needs, poverty, and limited access to health care, education and employment are major challenges for FSWs (16)(17). At the health facility level, while trust in certain healthcare providers can positively impact PrEP use for FSWs (18), persistent barriers exist owing to healthcare providers’ limited awareness of PrEP (19).

Rwanda has made significant strides in expanding access to PrEP, reflecting its commitment to reducing new HIV infections and improving the health of KPs, including FSWS. However, FSWs in the city of Kigali remains at high risk for new HIV infections(6). One study reported that only one-third of the participants consistently used condoms in the past month(5). Despite the potential benefits of PrEP in reducing new HIV infections(20), data on the level of awareness and willingness to use PrEP among FSWs in Rwanda are limited. To address this limitation, we conducted a survey among FSWs in Kigali to assess their awareness of and willingness to use PrEP. The findings from this research provide valuable insights to inform targeted interventions and support the expansion of PrEP services for FSWs in Rwanda.

## Methods

### Study design, setting, and population

This was a cross-sectional study that assessed awareness of and willingness to use PrEP among FSWs in the city of Kigali, Rwanda, between April and September 2024. Rwanda has a population of approximately 13 million (21), with an estimated 8,328--22,806 FSWS, which is predominantly based in Kigali(22). The annual HIV incidence is notably elevated for FSWS at 1.36%(6). The Rwandan guidelines for HIV prevention, treatment and care prioritize KPs for HIV prevention, including FSWS, who are eligible for PrEP if they are HIV negative, are over 18 years of age, and meet specific health criteria(23). PrEP is provided as oral tenofovir disoproxil fumarate-emtricitabine, with clinical evaluations and regular follow-ups to assess adherence and health(23). As of June 2023, the number of FSWS enrollees on PrEP has gradually increased to 10,789(24).

This study focused on FSWs in major local epidemic areas of Kigali, specifically communities with significantly higher rates of HIV prevalence or incidence than the national average(2)(6). These sites included Nyamirambo in the Nyarugenge district, a densely populated area known for its vibrant community and higher HIV rates among general adults(6); Remera Gisementi in the Gasabo district, characterized by nightlife and entertainment venues that may increase HIV transmission risk; and Kabuga in the Kicukiro district, which contains several neighborhoods with significant numbers of FSWS, contributing to elevated HIV prevalence rates.

## Data collection

We collaborated with FSWs associations in the neighborhoods of each study site, engaging FSWs who had previously worked with nongovernmental organizations (NGOs) experienced in health promotion. Each site had a designated head of association and an FSWS known for collaborating with other NGOs. Study participant recruitment took place three times a week—on Friday, Saturday, and Sunday evenings—during times recommended by the heads of FSWs associations, when most peers were present at the site. Only FSWs who presented at the study site for reasons unrelated to the study and were willing to speak with the selected heads of FSWs associations were recruited to participate if potentially eligible. Prior to data collection, eligible participants were required to provide written consent. Data collection was conducted by an experienced researcher and study assistant, both of whom are public health specialists. The participants who completed the survey received a cash incentive of 5,000 Rwandan francs (approximately $3.50) to compensate for opportunity costs and transportation expenses. The survey lasted between 1 and 2 hours per participant. Additionally, the heads of FSWs associations helped create a conducive environment for efficient and convenient data collection. To ensure quality assurance, weekly meetings were held with data collectors, the study coordinator, and the co-investigator to address data collection challenges, discuss solutions, and review the collected paper-based data before the survey forms were handed over to the data entry team.

## Study variables and measurements

The primary objective of the study was to assess the following: Awareness of PrEP: Defined as participants responding “yes” to the question, “Before today, have you ever heard of HIV-uninfected people taking ARV every day to reduce the risk of getting HIV?”. Willingness to use PrEP: Defined as participants indicating their willingness to “take ARV every day to lower the chances of contracting HIV.” Additional variables included the following: sociodemographic information, data on participants’ age, marital status, main activity, and highest level of education. Sexual history and HIV risk: Information gathered included the time since the last HIV test, the time since sex work started, the number of sexual partners in the seven days before the survey, and the frequency of condom use. STI screening and family planning methods: Collected data on whether participants had been screened for STIs in the past 12 months and the use of family planning methods. Table 4 shows the structure of the survey questions and the categorization of the variables.

## Sampling methods and size

The recent reported size estimate of FSWs in the country, with a median of 13,716, was not stratified by province or by district(22). To determine the sample for this study, we computed a sample size using an independent population. Given that this was a survey, our confidence interval was set at 95%, corresponding to a Z score of 1.96 and a margin of error of 5%. With this, given Z = 1.960, P = 0.5, and M = 0.05

The sample size formula was S= Z^2^ × P × [(1-P)/M ^2^].

S = (1.960)^2^ × 0.5 × [(1-0.5)/0.05^2^] = (3.8416 × 0.25)/0.0025

S= 384.16∼384 were equally distributed across the three study areas.

## Data analysis

The data were analyzed via IBM SPSS Statistics (version 21). Frequencies and proportions were calculated for categorical variables, whereas means or medians were used for continuous variables. For the outcome variable PrEP awareness, responses were dichotomized as ‘Yes’ (’Yes, I have heard of PrEP,’ ‘Yes, I know what PrEP is’) or ‘No’ (’No, I have never heard of PrEP,’ ‘No, I do not know what PrEP is’). For willingness to use PrEP, responses were dichotomized as ‘Yes’ or ‘No’ (’No, I do not know’). The mean age, median duration as an FSWS, and number of sexual partners in the previous seven days were also calculated.

Bivariate logistic regression, accounting for the number of respondents to the awareness and willingness questions, was used to examine associations between individual characteristics and these outcomes. The results are reported as unadjusted odds ratios (ORs) with 95% confidence intervals (CIs). The multivariable logistic regression model included individual characteristics that were significantly associated with the outcomes (p < 0.05) in the bivariate analysis. Associations are reported as adjusted odds ratios (aORs) with 95% CIs at a significance level of 0.05 for PrEP awareness and willingness to use.

## Ethical considerations

This study was approved by the Rwanda National Ethics Committee (RNEC 76/2024), which also approved the informed consent forms.

## Informed consent/Assent form

Written informed consent was obtained from all participants prior to their inclusion in the study. Participants were provided with detailed information about the study objectives, procedures, potential risks, and benefits. They were assured of their right to withdraw at any time without any consequences. Confidentiality of all personal data was guaranteed throughout the study.

## RESULTS

Among the 333 participants in this study (refer to data flow figure 1), the majority of participants were single (67%), unemployed in the formal workforce (66%), and held a primary school-level education (49.5%), with a mean age of 30 years (Table 1). Approximately 81% (258) of the sample reported having had an HIV test within six months prior to the survey, 59% reported being screened for an STI in the last 12 months, and 50% (165) had been engaged in sex work for 2–5 years. The median number of sexual partners in the last seven days was 7, with approximately 40% (132) reporting fewer than 5 partners during that period, approximately 49% (162) indicating the use of condoms sometime in the past week, and 72.5% indicating that they were using hormonal contraception at the time of the survey.

**Figure 1.**
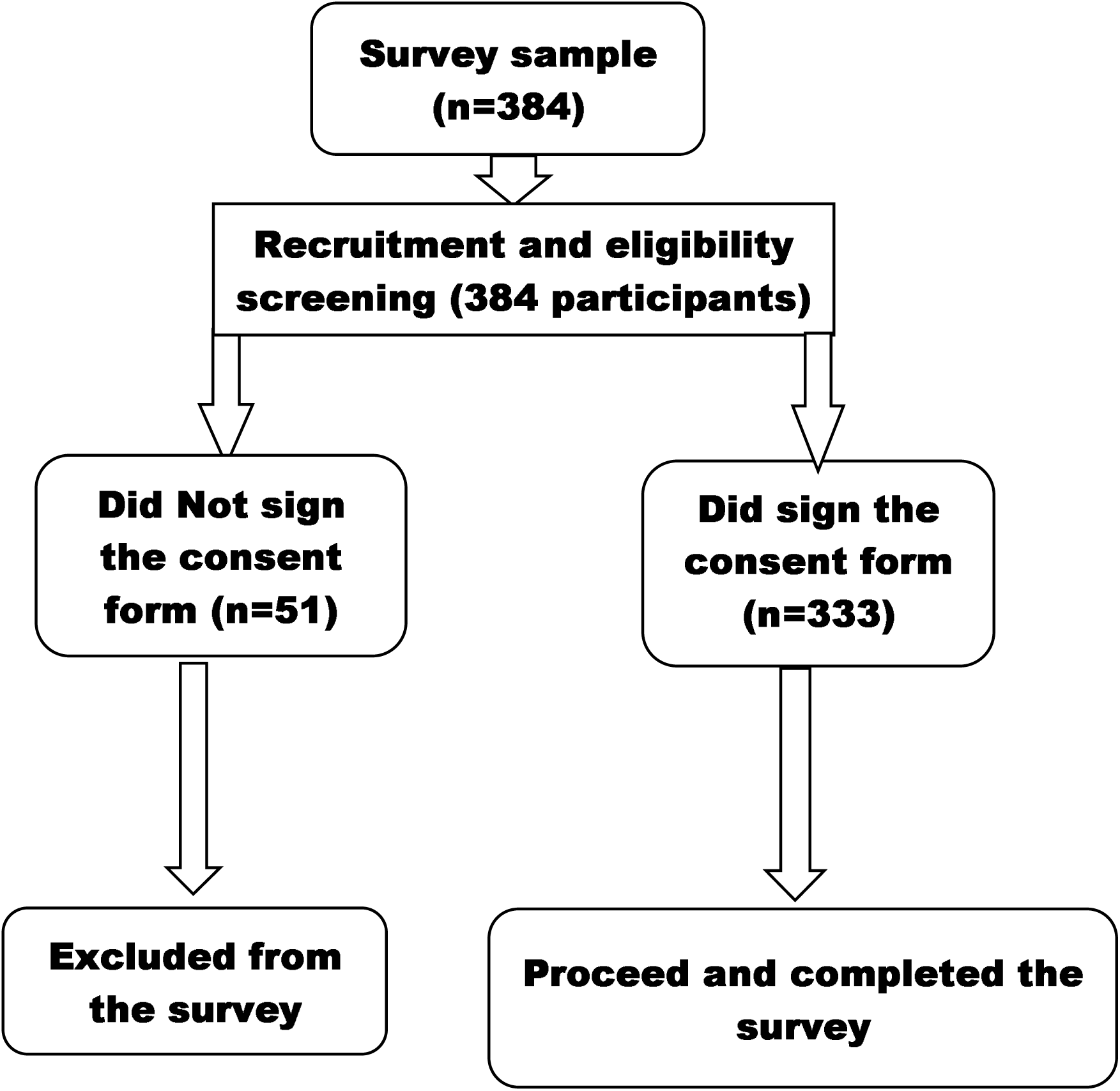
Participant flow in the study survey.

**Table 1.**
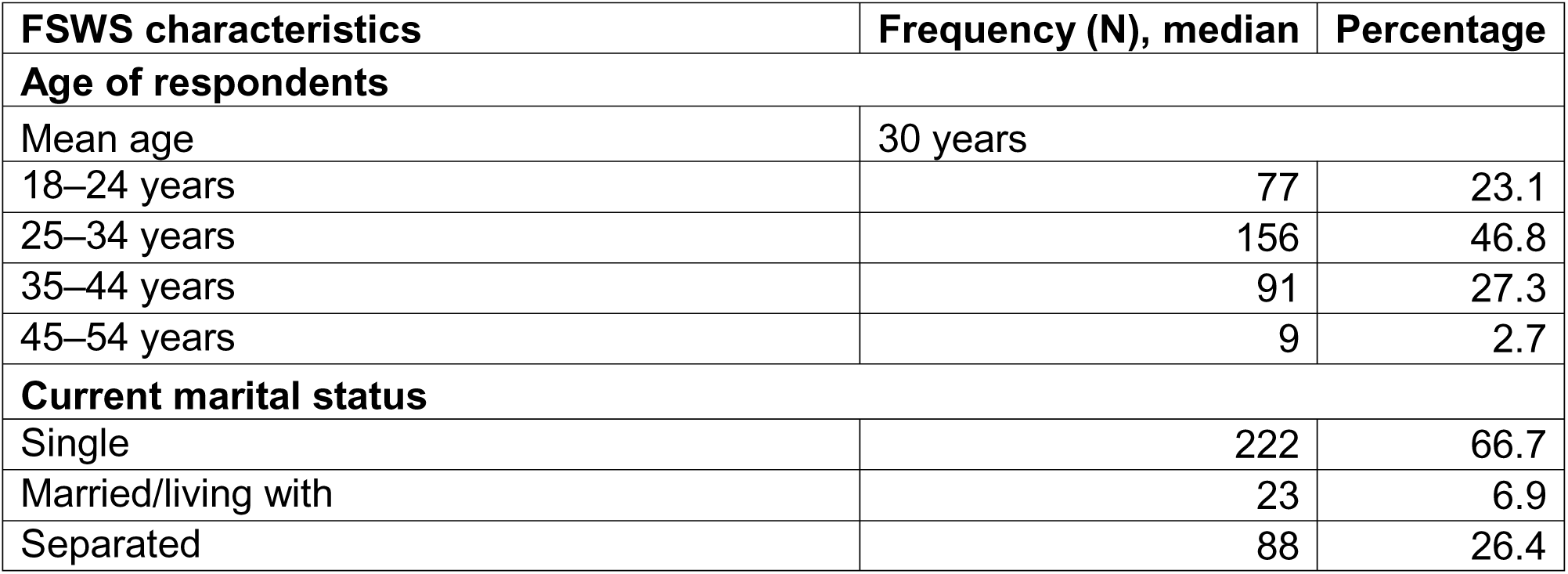
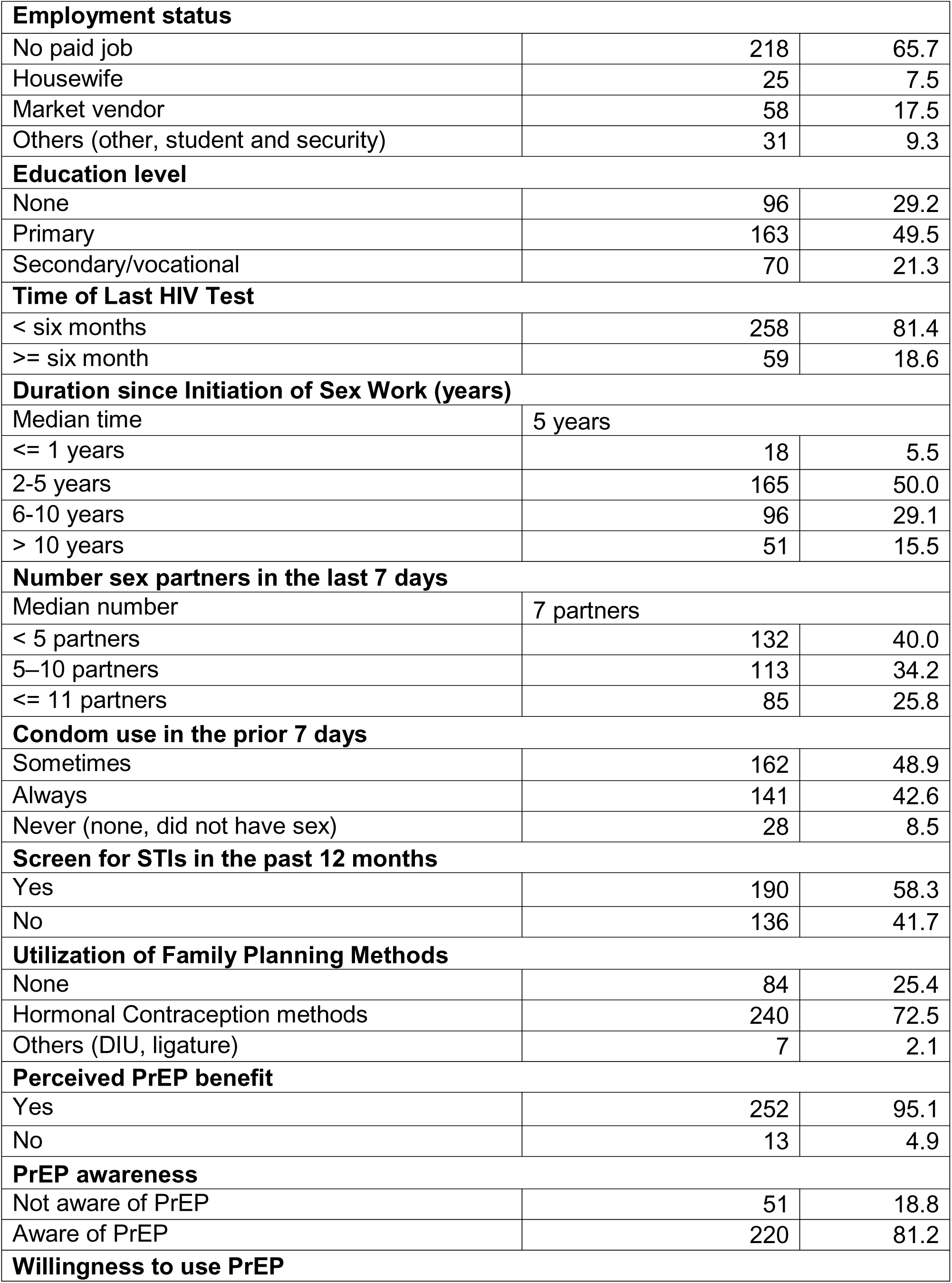
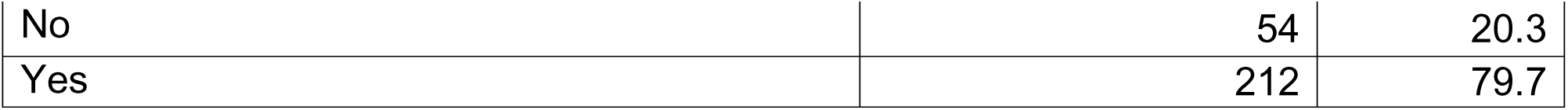
Survey participant characteristics.

| FSWS characteristics | Frequency (N), median |  | Percentage |
| --- | --- | --- | --- |
| Age of respondents |  |  |  |
| Mean age | 30 years |  |  |
| 18–24 years | 77 |  | 23.1 |
| 25–34 years | 156 |  | 46.8 |
| 35–44 years | 91 |  | 27.3 |
| 45–54 years | 9 |  | 2.7 |
| Current marital status |  |  |  |
| Single | 222 |  | 66.7 |
| Married/living with | 23 |  | 6.9 |
| Separated | 88 |  | 26.4 |

|  |  |  |
| --- | --- | --- |
| **Employment status** |  |  |
| No paid job | 218 | 65.7 |
| Housewife | 25 | 7.5 |
| Market vendor | 58 | 17.5 |
| Others (other, student and security) | 31 | 9.3 |
| **Education level** |  |  |
| None | 96 | 29.2 |
| Primary | 163 | 49.5 |
| Secondary/vocational | 70 | 21.3 |
| **Time of Last HIV Test** |  |  |
| < six months | 258 | 81.4 |
| >= six month | 59 | 18.6 |
| **Duration since Initiation of Sex Work (years)** |  |  |
| Median time | 5 years |  |
| <= 1 years | 18 | 5.5 |
| 2-5 years | 165 | 50.0 |
| 6-10 years | 96 | 29.1 |
| > 10 years | 51 | 15.5 |
| **Number sex partners in the last 7 days** |  |  |
| Median number | 7 partners |  |
| < 5 partners | 132 | 40.0 |
| 5–10 partners | 113 | 34.2 |
| <= 11 partners | 85 | 25.8 |
| **Condom use in the prior 7 days** |  |  |
| Sometimes | 162 | 48.9 |
| Always | 141 | 42.6 |
| Never (none, did not have sex) | 28 | 8.5 |
| **Screen for STIs in the past 12 months** |  |  |
| Yes | 190 | 58.3 |
| No | 136 | 41.7 |
| **Utilization of Family Planning Methods** |  |  |
| None | 84 | 25.4 |
| Hormonal Contraception methods | 240 | 72.5 |
| Others (DIU, ligature) | 7 | 2.1 |
| **Perceived PrEP benefit** |  |  |
| Yes | 252 | 95.1 |
| No | 13 | 4.9 |
| **PrEP awareness** |  |  |
| Not aware of PrEP | 51 | 18.8 |
| Aware of PrEP | 220 | 81.2 |
| **Willingness to use PrEP** |  |  |
| No | 54 | 20.3 |
| Yes | 212 | 79.7 |

### Awareness of and willingness to use PrEP

There was a high level of reported awareness of PrEP (81%) and the benefit of using PrEP to reduce the risk of HIV (95%) among the sample. In the bivariate analysis, PrEP awareness was significantly associated with condom use in the prior seven days, STI screening in the past 12 months, and perceived PrEP benefit (Table 2).

**Table 2.**
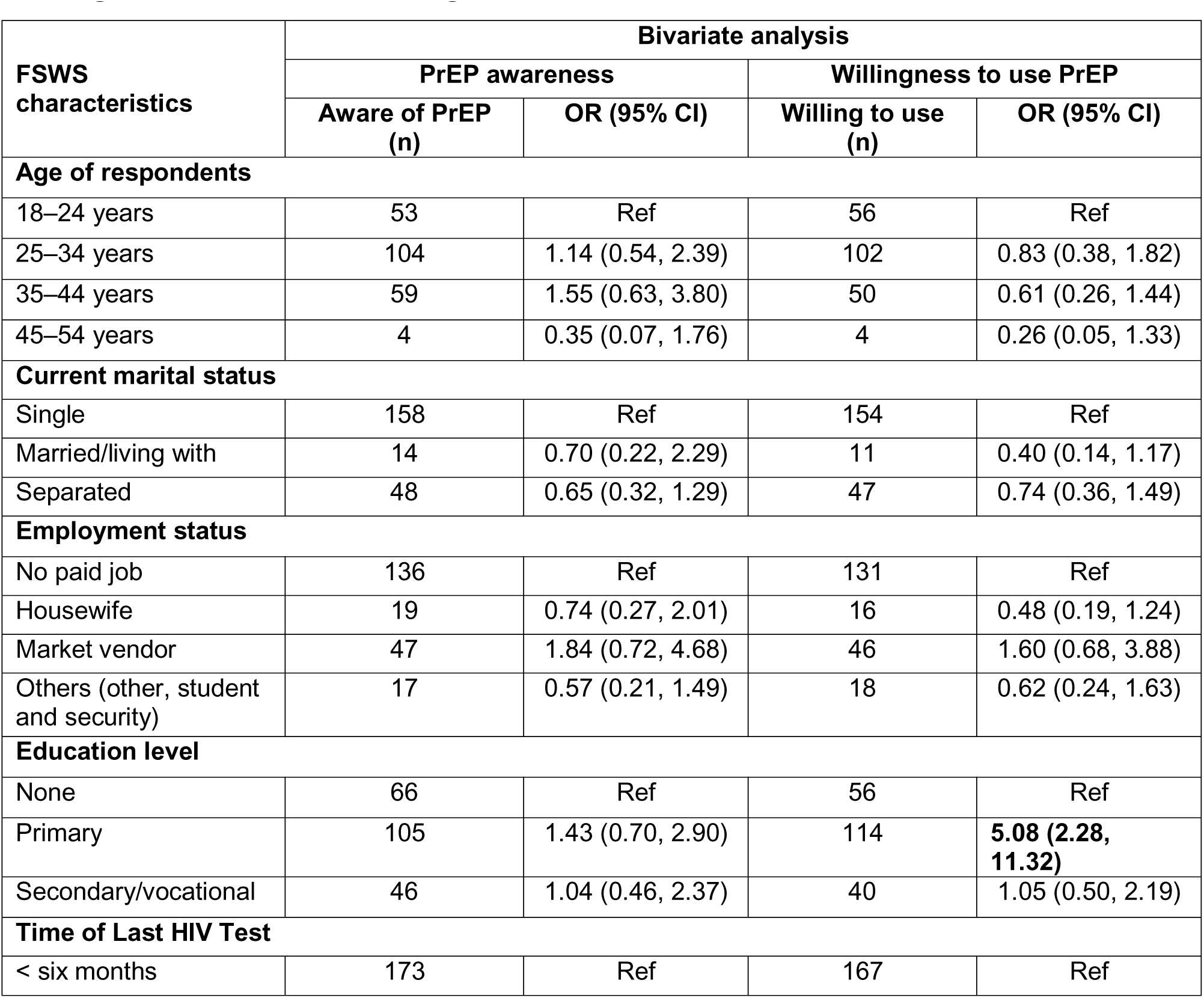
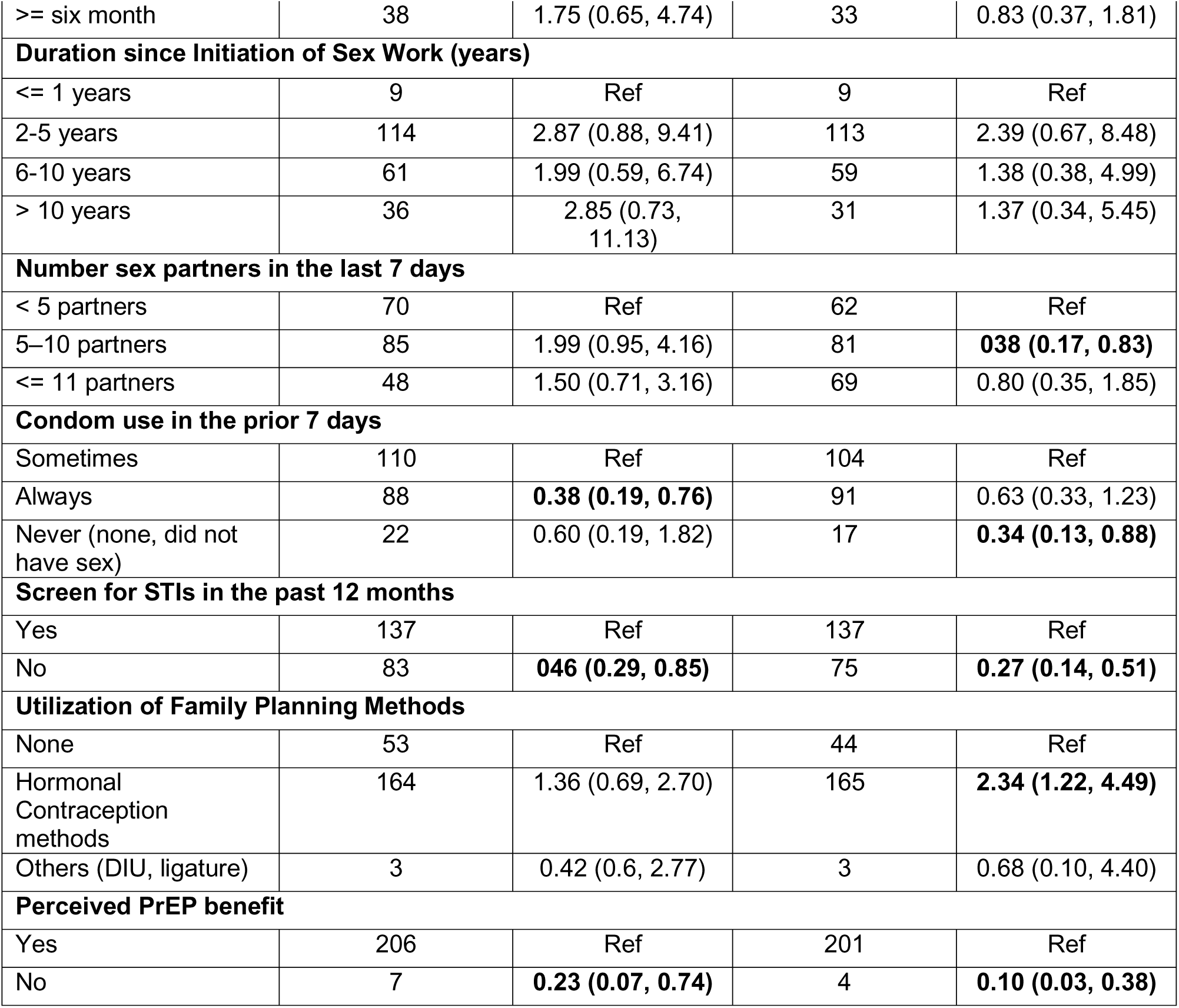
Bivariate analysis of survey characteristics and awareness of and willingness to use PrEP among FSWs.

According to the multivariable logistic regression analysis (Table 3), FSWs who reported always using condoms during sex in the prior seven days were less likely to be aware of PrEP (aOR: 0.40, 95% CI: 0.19–0.83) than those who sometimes used condoms. Similarly, FSWs who had not been screened for STIs in the past 12 months were less likely to be aware of PrEP (aOR: 0.43, 95% CI: 0.22–0.85) than those who had been screened. Additionally, FSWs who did not perceive a benefit from PrEP were less likely to be aware of it (aOR: 0.20, 95% CI: 0.05–0.67) than those who recognized its benefit.

**Table 3.** Multivariable Analysis of Survey Participants’ Characteristics and Awareness of, and Willingness to Use, PrEP.

| FSWS characteristics | Multivariable analysis |  |  |  |
| --- | --- | --- | --- | --- |
|  | PrEP awareness |  | Willingness to use PrEP |  |
|  | Aware of PrEP (n) | aOR (95% CI) | Willing to use (n) | aOR (95% CI) |
| **Education level** |  |  |  |  |
| None |  |  | 56 | Ref |
| Primary |  |  | 114 | **4.09 (1.62, 10.33)** |
| Secondary/vocational |  |  | 40 | 0.76 (0.30, 1.92) |
| **Number sex partners in the last 7 days** |  |  |  |  |
| < 5 partners |  |  | 62 | Ref |
| 5–10 partners |  |  | 81 | 1.46 (0.62, 3.45) |
| <= 11 partners |  |  | 69 | 1.27 (0.46, 3.49) |
| **Condom use in the prior 7 days** |  |  |  |  |
| Sometimes | 110 | Ref | 104 | Ref |
| Always | 88 | **0.40 (0.19, 0.83)** | 91 | 0.80 (0.35, 1.82) |
| Never (none, did not have sex) | 22 | 0.82 (0.25, 2.72) | 17 | 0.95 (0.26, 3.44) |
| **Screen for STIs in the past 12 months** |  |  |  |  |
| Yes | 137 | Ref | 137 | Ref |
| No | 83 | **0.43 (0.22, 0.85)** | 75 | **0.28 (0.12, 0.62)** |
| **Utilization of Family Planning Methods** |  |  |  |  |
| None |  |  | 44 | Ref |
| Hormonal Contraception methods |  |  | 165 | 1.53 (0.70, 3.32) |
| Others (DIU, ligature) |  |  | 3 | 1.38 (0.13, 14.56) |

| Perceived PrEP benefit |  |  |  |  |
| --- | --- | --- | --- | --- |
| Yes | 206 | Ref | 201 | Ref |
| No | 7 | **0.20 (0.05, 0.67)** | 4 | **0.06 (0.16, 0.29)** |

**Table 4.** Survey questions and variable categorizations.

| **Number** | **Survey questions** | **Categorization** |
| --- | --- | --- |
| 1 | **What is your age?** | 1.18–24 years<br>2.25–34 years<br>3.35–44 years<br>4.45–54 years |
| 2 | **What is your marital status?** | 1. Single<br>2. Married/living with<br>3. Separated |
| 3 | **What is your main activities?** | 1. No paid job<br>2. Housewife<br>3. Market vendor<br>4. Others (other, student and security) |
| 4 | **What is your highest level of education?** | 1. None<br>2. Primary<br>3. Secondary/vocational |
| 5 | **When is your last HIV test?** | 1. < six months<br>2. >= six month |
| 6 | **Since when did you start being sex workers?** | 1. <= 1 years<br>2. 2-5 years<br>3. 6-10 years<br>4. > 10 years |
| 7 | **How many partner have you had vaginal sex in the past seven days and past 12 months**<br>... 7 days | 1. < 5 partners<br>2. 5–10 partners<br>3. 11–20 partners<br>4. > 20 partners |
| 8 | **How often are condoms used during vaginal sex for the last seven days?** | 1. Never (none, did not have sex)<br>2. Always<br>3. Sometimes |
| 9 | **Do you use any method to delay or avoid pregnancy?** | 1. None<br>2. Hormonal (pills. implant, injectable) |
|  |  | 3. Others (DIU, ligature) |
| 10 | **Have you ever screened for STIs in the past 12 months?** | 1. Yes (screened)<br>2. Not screened |
| 11. | **Before today, how much would you say you knew about PrEP?** | 0. Not aware of PrEP<br>1. Aware of PrEP (all yes) |
| 12 | **Based on this introduction, do you think you could benefit from PrEP?** | 1. Yes<br>2. No |
| 13 | **PrEP is now available free of charge at health centers. Would you be willing to start taking PrEP in the next month to protect yourself against HIV?** | 0. Not willing to use (no, and I do not know)<br>1. Willing to use (all yes) |

Among FSWs respondents, 80% (212) reported being willing to use PrEP in the next month to protect themselves against HIV. In the bivariate analysis, willingness to use PrEP was significantly associated with education level, condom use in the previous seven days, STI screening in the previous 12 months, the number of sexual partners in the previous seven days, the use of family planning methods, and perceived PrEP benefits (Table 2). According to the multivariable logistic regression model, FSWs with a primary education level were more likely to be willing to use PrEP (aOR: 4.09, 95% CI: 1.62–10.33) than were those with no formal education. Additionally, FSWs who had not been screened for STIs in the past 12 months were less likely to be willing to use PrEP (aOR: 0.28, 95% CI: 0.12–0.62) than those who had been screened. Furthermore, FSWs who did not perceive a benefit from PrEP were less likely to be willing to use it (aOR: 0.06, 95% CI: 0.16–0.29) than those who recognized its benefit.

## DISCUSSION

In this study, we assessed awareness of and willingness to use PrEP among FSWs in three major HIV epidemic areas of Kigali. We found high levels of awareness and willingness to use within this population.

High awareness of PrEP among FSWs has been reported elsewhere in SSA. For example, a cohort of 700 HIV-negative FSWs in Dar es Salaam reported a PrEP awareness rate of 67% at enrollment, which increased to 97% after 12 months(8). Similarly, in Nigeria, a cross-sectional study of 344 FSWs receiving health promotion and prevention services at the One Stop Shop (OSS) reported a PrEP awareness rate of 76%(25), whereas an online survey reported a PrEP awareness rate of 95%(26).

Data on PrEP awareness among FSWs in Rwanda are limited. One of our studies reported 62% awareness among FSWs(27), whereas a study in Nigeria reported lower awareness; for example, a cross-sectional study in Anambra State reported an awareness rate of only 31%(28). These differences may be attributed to our convenience sampling of FSWs from major HIV epidemic areas in Kigali. This study was conducted two years later, reporting 62% PrEP awareness among FSWs(27). While the previous study used routine clinical data from a primary health facility, our study collected data from major HIV epidemic areas where FSWs networks are likely stronger. In contrast, the Anambra study involved brothel FSWs recruited through the snowball technique. Notably, our prior study assessing PrEP awareness among men who have sex with men (MSM) in Rwanda—a KP also eligible for PrEP—reported similarly high awareness levels to our findings(29), suggesting that access to PrEP information may be consistent across KPs in the country.

Awareness of PrEP was lower among FSWs who reported always using condoms than among those who used them less frequently, those who had never been screened for STIs in the past 12 months than among those who had, and those who did not perceive the benefits of PrEP compared with those who did. While these findings indicate high awareness among FSWs, they also highlight the need for targeted educational interventions to increase PrEP awareness within specific subgroups. Our findings indicate that FSWs who reported always using condoms had lower awareness of PrEP than did those who did not. This may suggest that condom users believe that their risk of HIV transmission is adequately reduced by condom use alone, resulting in low PrEP usage as an additional preventive measure. Public health programs should address the misconception that PrEP is only for those who use condoms inconsistently. Messaging should emphasize that PrEP provides an additional layer of protection even for those who consistently use condoms.

Furthermore, our findings revealed that FSWs who did not perceive any benefit from PrEP were significantly less likely to be aware of it. This highlights the crucial role of perceived benefits in shaping health behavior. Those who do not view PrEP as useful may not seek information or education about it, leading to lower levels of awareness. This lack of perceived benefit could stem from a misunderstanding of how PrEP works, a belief that other preventive measures (such as condoms) are sufficient, or misinformation about the effectiveness or side effects of PrEP. Without understanding the value that PrEP can provide in preventing HIV, FSWs may not engage with or prioritize information about it, resulting in lower awareness.

Our survey findings indicated a high willingness to use PrEP among FSWs, particularly among those with a primary education level, compared with those with no formal education. Furthermore, FSWs who had not undergone STI screening in the past 12 months were significantly less likely to be willing to use PrEP, as were those who did not perceive a benefit from it. These results emphasize the critical role of education, STI screening, and the perception of the benefits of PrEP in influencing the willingness to adopt PrEP among this population. Although no studies on PrEP willingness among FSWs in Rwanda have been conducted, previous research among MSM in the country reported a high willingness to use PrEP, exceeding 80%(9)(29).

Additionally, our findings showed that FSWs who had not been screened for STIs in the past 12 months were less likely to be willing to use PrEP. Other studies have reported that STI screening is associated with increased odds of willingness to use PrEP(30)(31), reinforcing the idea that regular health visits increase exposure to valuable health information. These findings suggest that regular STI screenings may provide additional health information, including information on HIV prevention methods such as PrEP. Those who were not screened may have avoided health facilities, lacked interest in PrEP information, or relied on other HIV prevention methods, such as condoms.

The high willingness to use PrEP reported in our findings suggests promising potential for PrEP uptake in this population, which could help reduce HIV acquisition in high-risk areas. A scoping review of PrEP adherence reported that the success of PrEP programs depends on willingness to use and prior awareness(32). Therefore, the high reported willingness to use PrEP among FSWs concentrated in major HIV epidemic areas in Kigali suggests a greater likelihood of PrEP uptake in these populations.

Similar findings have been reported across SSA, where high levels of willingness to use PrEP were observed. For example, in a cohort study conducted in Tanzania, 98% of the respondents were willing to use PrEP at enrollment, whereas 96% remained willing after 12 months(8). In another study in Anambra State, Nigeria and Uganda, 91% of respondents expressed a strong willingness to use PrEP (28)(33). Furthermore, a study in Ghana reported a willingness to use PrEP rate of 80% among FSWs(30). These results demonstrate a consistent trend of high acceptance and willingness to adopt PrEP in different regions of SSA.

Our study revealed that FSWs with a primary education level were more likely to express willingness to use PrEP than were those with no education. This highlights the potential influence of education on PrEP uptake and the need for targeted interventions in less educated populations. A study among adolescent girls in Rwanda has indicated an association between education level and general knowledge of HIV infection(34). Although this study did not focus specifically on FSWs, its findings suggest that higher education levels are associated with increased odds of having comprehensive HIV knowledge, including preventive measures. These results align with our findings, where FSWs with at least one primary education level were more willing to use PrEP. Similar studies, such as one in Nigeria(28), showed that FSWs seeking more HIV knowledge had higher odds of PrEP willingness, and research in Uganda also reported that education increases the willingness to use PrEP(33).

An emerging body of literature demonstrates that increasing PrEP awareness among FSWS is a key predictor of higher uptake and sustained long-term engagement with the intervention(8)(35).

Our findings showed that FSWs who did not perceive any benefit from PrEP were less likely to be willing to use it. This underscores the need for targeted education and awareness campaigns that emphasize the effectiveness of PrEP in preventing HIV, particularly among high-risk populations. This suggests the need to integrate comprehensive PrEP education into existing sexual health services for FSWs while ensuring that healthcare providers are trained to communicate the benefits of PrEP effectively. Additionally, addressing misconceptions and barriers to understanding the value of PrEP could significantly increase its uptake among FSWs.

## Limitations

The findings from this survey have several limitations. As a cross-sectional study, it could not report changes in awareness or willingness to use PrEP over time, which is essential for predicting significant uptake. Future studies could follow up on this assessment.

Additionally, the survey included FSWs who self-reported not currently using PrEP; however, it did not ask whether they had used it before, nor could it verify the accuracy of these self-reports.

Furthermore, the survey was conducted during a period when the PrEP rollout project was being implemented across most primary health facilities in Rwanda. This may have resulted in more information being available to eligible individuals, including FSWs, contributing to the observed high awareness of PrEP.

Although the target sample included 384 participants, only 333 participants were analyzed because of incomplete responses, missing data, and unmet inclusion criteria. Nonetheless, the analyzed sample remains sufficient for meaningful analysis.

Finally, the survey focused on major HIV epidemic areas in Kigali, which limits the generalizability of the findings to other regions in the country.

Despite these limitations, the survey has notable strengths. There are limited data on PrEP awareness and willingness to use it among FSWs in Rwanda, and the findings provide baseline evidence-based information.

## Conclusion

The survey findings highlighted high awareness and willingness to use PrEP among FSWs in Rwanda, suggesting that they may become increasingly receptive to its adoption as the implementation project progresses. However, FSWs who consistently use condoms should receive targeted HIV prevention information that complements condom use, including PrEP. Additionally, enhancing awareness among those with limited STI screening and low perceived benefits is crucial. Emphasizing education and regular health screenings can further foster acceptance of PrEP. Organizing outreach campaigns focused on STI screening and PrEP benefits will encourage FSWS to engage in regular check-ups and gain essential health information related to HIV risk and PrEP’s advantages.

## Conflict of interest

The authors declare that they have no conflicts of interest.

## Authors’ contributions

**AN** and **AM** contributed significantly to the study protocol development and data collection. **AM** conducted the data analysis. **AN** led the manuscript drafting, with substantial input from **AM** on design and editing. **AM** also revised the work. All authors approved the final version of the manuscript.

## Data Availability

All data produced in the present study are available upon request to the lead or senior author

## Acknowledgment

We acknowledge the heads of FSWS associations from the study sites for their support in participant recruitment and for providing convenient locations for the survey. Special thanks to King Faisal Hospital for offering early career researcher grant opportunities.

## Funding

This research study was supported by King Faisal Hospital, Rwanda, under Grant Award Ref: KFH/507/23/CEO/RN.

## Notes

### Competing Interest Statement

The authors have declared no competing interest.

### Author Declarations

This study was approved by the Rwanda National Ethics Committee (RNEC 76/2024), which also approved the informed consent forms. Written informed consent was obtained from all participants prior to their inclusion in the study. Participants were provided with detailed information about the study objectives, procedures, potential risks, and benefits. They were assured of their right to withdraw at any time without any consequences. Confidentiality of all personal data was guaranteed throughout the study.

## References

1. UNAIDS. Global HIV statistics:FACT SHEET 2024 [Internet]. The Joint United Nations Programme on HIV/AIDS. 2024 [cited 2024 Aug 22]. p. 3. Available from: https://www.unaids.org/sites/default/files/media_asset/UNAIDS_FactSheet_en.pdf

2. UNAIDS. IN DANGER: UNAIDS Global AIDS Update 2022. Geneva: Joint United Nations Programme on HIV/ AIDS; 2022. Licence: CC BY-NC-SA 3.0 IGO. [Internet]. 2022. Available from: https://www.unaids.org/sites/default/files/media_asset/2022-global-aids-update_en.pdf

3. Impact PHI V, Rphia A. February 2020 Rwanda Population-Based Hiv Impact Assessment. (February 2020):2–8.

4. Nsanzimana S, Mills EJ, Harari O, Mugwaneza P, Karita E, Uwizihiwe JP, Park JJ, Dron L, Condo J, Bucher H TK. Prevalence and incidence of HIV among female sex workers and their clients: modelling the potential effects of intervention in Rwanda. BMJ Glob Heal. 2020;5(8):e002300.

5. Ingabire R, Parker R, Nyombayire J, Ko JE, Mukamuyango J, Bizimana J, et al. Female sex workers in Kigali, Rwanda: a key population at risk of HIV, sexually transmitted infections, and unplanned pregnancy. Int J STD & AIDS [Internet]. 2019 May;30(6):557—568. Available from: https://europepmc.org/articles/PMC6512058

6. Nsanzimana S, Rwibasira GN, Malamba SS, Musengimana G, Kayirangwa E, Jonnalagadda S, et al. HIV incidence and prevalence among adults aged 15-64 years in Rwanda: Results from the Rwanda Population-based HIV Impact Assessment (RPHIA) and District-level Modeling, 2019. Int J Infect Dis IJID Off Publ Int Soc Infect Dis. 2022 Mar;116:245–54.

7. CDC. Effectiveness of Prevention Strategies to Reduce the Risk of Acquiring or Transmitting HIV [Internet]. Center for Disease Control and prevention. [cited 2023 Oct 2]. p. 1. Available from: https://www.cdc.gov/hiv/risk/estimates/preventionstrategies.html#anchor_1562942347

8. Faini D, Munseri P, Sandstrom E, Hanson C, Bakari M. Awareness, Willingness and Use of HIV Pre-Exposure Prophylaxis Among Female Sex Workers Living in Dar-es-Salaam, Tanzania. AIDS Behav. 2023 Jan;27(1):335–43.

9. Munyaneza A, Adedimeji A, Kim H-Y, Shi Q, Hoover DR, Ross J, et al. Awareness and Willingness to Use HIV Pre-exposure Prophylaxis Among Men Who Have Sex With Men in Rwanda: A Cross-Sectional Descriptive Survey. J Assoc Nurses AIDS Care. 2021;32(6):693–700.

10. Kabaghe AN, Singano V, Payne D, Maida A, Nyirenda R, Mirkovic K, et al. Awareness of and willingness to use oral pre-exposure prophylaxis (PrEP) for HIV prevention among sexually active adults in Malawi: results from the 2020 Malawi population-based HIV impact assessment. BMC Infect Dis [Internet]. 2023;23(1):712. Available from: 10.1186/s12879-023-08683-1

11. Faini D, Munseri P, Sandstrom E, Hanson C, Bakari M. Awareness, Willingness and Use of HIV Pre-Exposure Prophylaxis Among Female Sex Workers Living in Dar-es-Salaam, Tanzania. AIDS Behav. 2023 Jan;27(1):335–343. doi: 10.1007/s10461-022-03769-4. Epub 2022.

12. Tomko C, Park JN, Allen ST, Glick J, Galai N, Decker MR, Footer KHA, Sherman SG. Awareness and Interest in HIV Pre-Exposure Prophylaxis Among Street-Based Female Sex Workers: Results from a US Context. AIDS Patient Care STDS. 2019 Feb;33(2):49–57. doi: 10.

13. Ghayda RA, Hong SH, Yang JW, Jeong GH, Lee KH, Kronbichler A, Solmi M, Stubbs B, Koyanagi A, Jacob L, Oh H, Kim JY, Shin JI, Smith L. A Review of Pre-Exposure Prophylaxis Adherence among Female Sex Workers. Yonsei Med J. 2020 May;61(5):349–358. doi: 10.33.

14. Van der Elst EM, Mbogua J, Operario D, Mutua G, Kuo C, Mugo P, et al. High acceptability of HIV pre-exposure prophylaxis but challenges in adherence and use: qualitative insights from a phase I trial of intermittent and daily PrEP in at-risk populations in Kenya. AIDS Behav. 2013 Jul;17(6):2162–72.

15. Aidsmap. Three forms of PrEP stigma in Kenya [Internet]. AIDSmap. 2019 [cited 2023 Dec 16]. p. 1. Available from: https://www.aidsmap.com/news/jul-2019/three-forms-prep-stigma-kenya

16. Emmanuel G, Folayan M, Undelikwe G, Ochonye B, Jayeoba T, Yusuf A, Aiwonodagbon B, Bilali C, Umoh P, Ojemeiri K, Kalaiwo A. Community perspectives on barriers and challenges to HIV pre-exposure prophylaxis access by men who have sex with men and female se.

17. Palar K, Wong MD, Cunningham WE. Competing subsistence needs are associated with retention in care and detectable viral load among people living with HIV. J HIV AIDS Soc Serv. 2018;17(3):163–79.

18. Beauchamp G, Donnell D, Hosek S, Anderson PL, Chan KCG, Dye BJ, et al. Trust in the provider and accurate self-reported PrEP adherence among adolescent girls and young women in South Africa and Zimbabwe: HPTN 082 study. BMC Womens Health [Internet]. 2023;23(1):276. Available from: 10.1186/s12905-023-02418-9

19. Kambutse I, Igiraneza G, Shenoi S, Ogbuagu O. Correction: Perceptions of HIV transmission and pre-exposure prophylaxis among health care workers and community members in Rwanda. PLoS One. 2019;14(2):e0212933.

20. PEPFAR. The US President’s Emergency Plan for AIDS Relief. PEPFAR 2021 Country and Regional Operational Plan (COP/ROP) Guidance for all PEPFAR Countries. 2021.

21. National Institute of Statistics of Rwanda (NISR); The Fifth Rwanda Population and Housing Census, Main Indicators Report, February 2023 [Internet]. Kigali; 2023. Available from: file:///C:/Users/hp/Downloads/RPHC5_MainIndicatorsReport_Final.pdf

22. Musengimana G, Tuyishime E, Remera E, Dong M, Sebuhoro D, Mulindabigwi A, et al. Female sex workers population size estimation in Rwanda using a three-source capture-recapture method. Epidemiol Infect. 2021 Mar;149:e84.

23. MoH. GUIDELINES FOR HIV PREVENTION, TREATMENT AND CARE IN RWANDA: Edition 2022 [Internet]. Ministry of Health, Rwanda Biomedical center. 2022 [cited 2023 Oct 3]. Available from: https://rbc.gov.rw/fileadmin/user_upload/guidelines 23/Final GUIDELINES FOR HIV PREVENTION%2C TREATMENT AND CARE IN RWANDA 2022 c.pdf

24. RBC. HIV,STIs, and Viral Hepatatis Program Annual Report 2022-2023 [Internet]. Kigali; 2023. Available from: https://www.rbc.gov.rw/fileadmin/user_upload/report23/HIV Annual report 2022–2023.pdf

25. Ughweroghene Kingston Omo-Emmanuel DCU, Osazee B, Airiagbonbu, Usman HB, Jegede FE, Aka-Okeke C, et al. Assessment of Awareness, Willingness, and Practice of Human Immunodeficiency Virus Pre-Exposure Prophylaxis Among Female Sex Workers in Uyo, Akwa Ibom, Nigeria. Texila Int J Public Heal. 2023;(2520–3134).

26. Emmanuel G, Folayan M, Undelikwe G, Ochonye B, Jayeoba T, Yusuf A, et al. Community perspectives on barriers and challenges to HIV pre-exposure prophylaxis access by men who have sex with men and female sex workers access in Nigeria. BMC Public Health. 2020;20(1):1–10.

27. Munyaneza A, Bhutada K, Shi Q, Zotova N, Nsereko E, Muhoza B, et al. High retention among key populations initiated on HIV pre-exposure prophylaxis in Kigali City, Rwanda. J Int AIDS Soc. 2024 Nov;27(11):e26392.

28. Nwagbo, E. C., Ekwunife, O. I., Mmeremikwu, A. C. A, Ojide CK. Awareness of and willingness to use pre-exposure prophylaxis to prevent HIV infection among female sex workers in Anambra State, south-eastern Nigeria. Afr J Clin Exper Microbiol. 2023;24(2):168–76.

29. Munyaneza A, Patel V V, Gutierrez NR, Shi Q, Muhoza B, Kubwimana G, et al. Awareness and willingness to use pre-exposure prophylaxis for HIV prevention among men who have sex with men in Rwanda: findings from a web-based survey. Front public Heal. 2024;12:1325029.

30. Guure C, Afagbedzi S, Torpey K. Willingness to take and ever use of pre-exposure prophylaxis among female sex workers in Ghana. Medicine (Baltimore). 2022 Feb;101(5):e28798.

31. Peng B, Yang X, Zhang Y, Dai J, Liang H, Zou Y, et al. Willingness to use pre-exposure prophylaxis for HIV prevention among female sex workers: a cross-sectional study in China. HIV AIDS (Auckl). 2012;4:149–58.

32. Ghayda RA, Hong SH, Yang JW, Jeong GH, Lee KH, Kronbichler A, et al. A Review of Pre-Exposure Prophylaxis Adherence among Female Sex Workers. Yonsei Med J. 2020 May;61(5):349–58.

33. Witte SS, Filippone P, Ssewamala FM, Nabunya P, Bahar OS, Mayo-Wilson LJ, et al. PrEP acceptability and initiation among women engaged in sex work in Uganda: Implications for HIV prevention. EClinicalMedicine. 2022 Feb;44:101278.

34. Kawuki J, Gatasi G, Sserwanja Q, Mukunya D, Musaba MW. Comprehensive knowledge about HIV/AIDS and associated factors among adolescent girls in Rwanda: a nationwide cross-sectional study. BMC Infect Dis. 2023 Jun;23(1):382.

35. Nunn AS, Brinkley-Rubinstein L, Oldenburg CE, Mayer KH, Mimiaga M, Patel R, et al. Defining the HIV pre-exposure prophylaxis care continuum. AIDS. 2017 Mar;31(5):731–4.

